# Validity and Reliability of the Executive Function Scale (UEF-1) in Cuban University Students

**DOI:** 10.1101/2024.06.07.24308560

**Authors:** Diego D. Díaz-Guerra, Marena de la C. Hernández-Lugo, Carlos Ramos-Galarza, Yunier Broche-Pérez

**Author notes:** Corresponding author information: Prof. Diego D. Díaz-Guerra Psychology Department, Universidad Central “Marta Abreu” de Las Villas Km 5 ½, Santa Clara, Villa Clara, Cuba, 54 830.

## Abstract

**Introduction:** Executive functions are higher cognitive skills involved in planning, organization, decision-making, impulse control, and working memory. It is essential to have tools that allow for the accurate and reliable assessment of this construct in university students. This study aims to evaluate the validity and reliability of the Executive Functions Scale for University Students (UEF-1) in the Cuban population.

**Methods:** A cross-sectional study was conducted in which an online survey was administered to 1092 Cuban university students representing 14 of the country’s 16 provinces. Descriptive analyses, confirmatory factor analyses, and Pearson correlation analyses were used to assess the psychometric properties of the scale.

**Results:** Significant correlations were obtained between the scale factors, and the original seven-factor structure was confirmed. The scale demonstrated good internal consistency and overall reliability (α = .91, ω = .91).

**Conclusions:** The study provided evidence that the UEF-1 is a reliable and valid tool for assessing executive functions in Cuban university students. This measure provides a comprehensive understanding of the cognitive abilities and functioning of Cuban university students, allowing for the identification of specific areas of executive functioning that may benefit from additional support or intervention.

## Introduction

Executive functions play a central role in academic performance, problem-solving, decision-making, and success in daily life (Drigas & Karyotaki, 2019; Ramos-Galarza et al., 2020; Reynolds et al., 2019; Zelazo & Carison, 2020). These higher cognitive skills enable individuals to regulate their behavior, maintain attention, inhibit automatic responses, switch tasks, and plan future actions (Cristofori et al., 2019; Doebel, 2020; Ramos-Galarza et al., 2022).

In the university context, where students are undergoing a period of significant transition and personal development, executive functions are especially relevant (Ramos-Galarza et al., 2020; Salas-Gomez et al., 2020). To successfully address the demands of the university environment, students need to have adequate executive functioning, allowing them to efficiently plan and organize tasks, maintain concentration and motivation, regulate their emotions, and adapt to new and changing situations (Cartwright et al., 2019; Hilton et al., 2022; Ramos-Galarza et al., 2020; Salas-Gomez et al., 2020).

Previous research has shown that students with better executive functioning tend to achieve higher grades, have greater satisfaction with their university experience, and experience less stress and emotional difficulties. Conversely, students with deficiencies in executive functions may struggle to manage their academic workload, encounter difficulties in decision-making and planning, and experience challenges in stress management and self-regulation (Hilton et al., 2022; Moriña & Biagiotti, 2022; Nieto et al., 2020; Pinochet-Quiroz et al., 2022; Ramos-Galarza et al., 2020; Reynolds et al., 2019; Zhang et al., 2019).

Given the importance of executive functions in the university setting, it is essential to have a reliable and valid assessment tool to measure these skills in the student population (Ramos-Galarza et al., 2020; Romero-Galisteo et al., 2022; Rowe et al., 2021). In this context, the University Executive Function Scale (UEF-1) has been developed as a potentially useful tool for assessing executive functioning in this population (Ramos-Galarza et al., 2023). This scale was designed and validated in Ecuadorian and Chilean populations where it demonstrated excellent validity and reliability results; however, no research has yet been reported on the evaluation of the psychometric properties of the UEF-1 in other Spanish-speaking contexts.

Furthermore, in the Cuban university population, there are no specific assessment tools for executive functions available. Instead, generic tests designed for other purposes but allowing clinical-level evaluation of executive functions have been used (e.g., Jiménez-Puig et al., 2019).

Therefore, this study aims to evaluate the psychometric properties of the *University Executive Function Scale* for university students in the Cuban population. This research will provide valuable information about the executive functioning of Cuban students and contribute to the development of specific interventions and support programs to enhance these skills in this population.

## Materials and methods

### Study Design and Participants

This research was based on a cross-sectional study. An online survey was conducted using Google Forms^®^ from January to March 2024, which was distributed through WhatsApp groups, Facebook, email lists, and websites. No incentives were offered for participation. All Cuban citizens over 18 years old who were pursuing university studies within the country were eligible to participate. A total of 1092 students (660 females and 432 males) with a mean age of 20.5 participated. The study included representation from 14 out of the 16 Cuban provinces.

### Measures

*Demographic Information*: Demographic variables explored included age, gender, and province where university studies were being pursued.

*University Executive Function Scale* (UEF-1): This scale was designed and validated for Spanish-speaking participants by Ramos-Galarza et al. (2023) in a sample of 1373 Chilean and Ecuadorian students. It consists of 31 items and measures 7 executive functions. The subscales it assesses are: Conscious Monitoring of Responsibilities (F1 items 2, 8, 9, 15, and 27), Supervisory Attention System (F2 items 10, 14, 22, 28, and 13), Conscious Regulation of Behavior (F3 items 3, 11, 16, 17, 18, and 19); Behavior Verification for Learning (F4 items 20, 23, 24, and 30); Decision Making (F5 items 5, 12, and 21), Conscious Regulation of Emotions (F6 items 4, 25, 29, and 31), and Task-solving Element Management (F7 items 1, 6, 7, and 26). Responses are provided on a 5-point Likert scale (1 = strongly disagree to 5 = strongly agree).

The original version of the scale was aimed at Spanish-speaking students, so it was not necessary to translate the items. However, following the recommendations of Fenn et al. (2020) for the adaptation and translation of questionnaires, the original version of the technique was examined by three linguistics specialists affiliated with the Department of Linguistics at the Universidad Central “Marta Abreu” de Las Villas. The experts concluded that the linguistic and grammatical structure of the scale was suitable for the Cuban context, therefore, no modifications were necessary in the scale. A pilot study was carried out, involving a sample of 50 students, to assess the suitability of the questions in relation to the language used within the Cuban context. The findings of the pilot study confirmed the recommendations put forth by the linguistic experts. These results support the use of the questionnaire in the Cuban population without the need for additional linguistic or grammatical modifications.

### Procedure and Data Analysis

Informed consent was obtained from all participants included in the study. The ethics committee of the Department of Psychology of the Universidad Central “Marta Abreu” de Las Villas approved the study protocol. All procedures performed in this study were following the ethical standards of the 1964 Helsinki Declaration.

The data were processed using the statistical software JASP version 0.18.3.0. Descriptive analyses were employed to understand the characteristics of the participants, and confirmatory factor analysis (CFA) was used to evaluate the factorial structure of the UEF-1. Pearson correlation analyses were conducted to assess the relationship between the executive functions comprising the instrument, with correlation indices of r ≤ .10 considered small, r = .20 considered moderate, and r ≥ .30 considered large (Funder & Ozer, 2019).

Acceptable values for the comparative fit index (CFI) and the Tucker-Lewis index (TLI) were assumed to be between .90 and .95 (Hu & Bentler, 1999). As for the root mean square error of approximation (RMSEA), values below .10 were considered acceptable (Xia & Yang, 2019). Cronbach’s α and McDonald’s ω were used to evaluate reliability, with values equal to or greater than .70 indicating good internal consistency (Dunn et al., 2013; Kelley, 2018; Shrestha, 2021).

To assess normality, the Kolmogorov-Smirnov test was employed. The p-values obtained were less than .05, indicating that the items of the UEF-1 were not normally distributed. Additionally, the analysis of skewness and kurtosis revealed values exceeding ±1, providing conclusive evidence that the data does not adhere to a normal distribution (Mishra et al., 2019). Considering the lack of normality in the sample, the weighted least squares mean, and variance adjusted estimator (WLSMV) were used for CFA (Xia & Yang, 2019).

## Results

### Characteristics of the sample

The sample consisted of 1092 university students, of which 39.6% were male and the remaining 60.4% were female, aged between 18 and 25 years old. Most participants belonged to the central region of the country (93%), particularly in the province of Villa Clara (76.8%) (Table 1).

**Table 1.** Participant demographic data (n=1092)

| Demographic variable | Mean ± SD or n (%) |
| --- | --- |
| Age | 20.05 ± 1.72 |
| Gender |  |
| Female | 432 (39.6) |
| Male | 660 (60.4) |
| Province |  |
| Occidental Region | 69 (6.4) |
| Pinar del Río | 1 (0.1) |
| Artemisa | 4 (0.4) |
| La Habana | 61 (5.6) |
| Isla de la Juventud | 1 (0.1) |
| Mayabeque | 1 (0.1) |
| Matanzas | 1 (0.1) |
| Central Region | 1016 (93.0) |
| Cienfuegos | 44 (4.0) |
| Villa Clara | 839 (76.8) |
| Sancti Spíritus | 38 (3.5) |
| Ciego de Ávila | 23 (2.1) |
| Camagüey | 72 (6.6) |
| Oriental Region | 7 (0.7) |
| Las Tunas | 4 (0.4) |
| Granma | 2 (0.2) |
| Santiago de Cuba | 1 (0.1) |

Note. SE (Standard Deviation)

Table 2 displays the values corresponding to the analysis of means, standard deviation, correlations between the total items, and their loadings. The items showed above-average approval rates and considerable variation.

**Table 2.** Descriptive statistics, item-test correlations (*r*), and loadings of the UEF-1 items.

| Items | $M$ | $SD$ | $r$ | Loadings |
| --- | --- | --- | --- | --- |
| F1 Conscious monitoring of responsibilities |  |  |  |  |
| UEF2 | 4.01 | 1.04 | .50 | .70 |
| UEF8 | 3.80 | 1.90 | .43 | .63 |
| UEF9 | 4.17 | .89 | .55 | .79 |
| UEF15 | 4.14 | .99 | .41 | .61 |
| UEF27 | 4.07 | 1.03 | .54 | .77 |
| F2 Supervisory Attention System |  |  |  |  |
| UEF10 | 3.60 | 1.21 | .60 | .77 |
| UEF14 | 3.96 | 1.07 | .59 | .74 |
| UEF22 | 3.70 | 1.11 | .63 | .81 |
| UEF28 | 3.47 | 1.26 | .57 | .73 |
| UEF13 | 3.35 | 1.21 | .59 | .74 |
| F3 Conscious regulation of behavior |  |  |  |  |
| UEF3 | 3.87 | 1.17 | .46 | .56 |
| UEF11 | 3.30 | 1.43 | .42 | .54 |
| UEF16 | 4.53 | .84 | .47 | .66 |
| UEF17 | 4.01 | 1.07 | .47 | .59 |
| UEF18 | 4.13 | 1.05 | .38 | .49 |
| UEF19 | 3.95 | 1.04 | .35 | .46 |
| F4 Verification of behavior to learn |  |  |  |  |
| UEF20 | 4.12 | 1.08 | .52 | .81 |
| UEF23 | 4.17 | 1.23 | .39 | .67 |
| UEF24 | 4.06 | 1.22 | .46 | .71 |
| UEF30 | 4.06 | 1.06 | .56 | .81 |
| F5 Decision making |  |  |  |  |
| UEF5 | 4.29 | .95 | .43 | .64 |
| UEF12 | 4.12 | .96 | .55 | .80 |
| UEF21 | 3.85 | 1.07 | .51 | .73 |
| F6 Conscious regulation of emotions |  |  |  |  |
| UEF4 | 3.65 | 1.17 | .49 | .77 |
| UEF25 | 3.65 | 1.20 | .45 | .69 |
| UEF29 | 3.66 | 1.29 | .53 | .82 |
| UEF31 | 3.78 | 1.20 | .50 | .84 |
| F7 Management of elements to solve tasks |  |  |  |  |
| UEF1 | 4.01 | 1.11 | .50 | .75 |
| UEF6 | 3.65 | 1.31 | .51 | .78 |
| UEF7 | 3.94 | 1.25 | .46 | .71 |
| UEF26 | 4.13 | 1.64 | .41 | .66 |

Note: $M$ = mean, $SD$ = standard deviation, $r$ = correlation index.

It was observed that all item-total correlations were acceptable. Similarly, all loadings were significant (p < .001). Furthermore, statistically significant correlations of moderate and large magnitudes between the factors of the scale were found (Table 3).

**Table 3.** Correlation matrix between executive functions.

|  | F 1 |  | F 2 |  | F 3 |  | F 4 |  | F 5 |  | F 6 |  | F 7 |
| --- | --- | --- | --- | --- | --- | --- | --- | --- | --- | --- | --- | --- | --- |
| F 1 | — |  |  |  |  |  |  |  |  |  |  |  |  |
| F 2 | 0.614 | *** | — |  |  |  |  |  |  |  |  |  |  |
| F 3 | 0.426 | *** | 0.546 | *** | — |  |  |  |  |  |  |  |  |
| F 4 | 0.511 | *** | 0.559 | *** | 0.427 | *** | — |  |  |  |  |  |  |
| F 5 | 0.473 | *** | 0.486 | *** | 0.475 | *** | 0.324 | *** | — |  |  |  |  |
| F 6 | 0.300 | *** | 0.443 | *** | 0.558 | *** | 0.235 | *** | 0.508 | *** | — |  |  |
| F 7 | 0.400 | *** | 0.435 | *** | 0.469 | *** | 0.446 | *** | 0.375 | *** | 0.357 | *** | — |

Note. F = Factor; \*\*\* = $p < .001$

The expected seven-factor solution showed satisfactory fit indices, χ2(413) = 1823.68, p < .001; RMSEA = .056, 90% CI [.053, .059], p < .001; CFI = .981; TLI = .979 (Figure 1). Thus, the initial solution of the seven proposed factors in the original scale is confirmed. It was not considered necessary to test a second-order model, considering a central factor of executive functions, as in the original validation of the scale, it was found that it does not present an acceptable fit.

**Figure 1.**
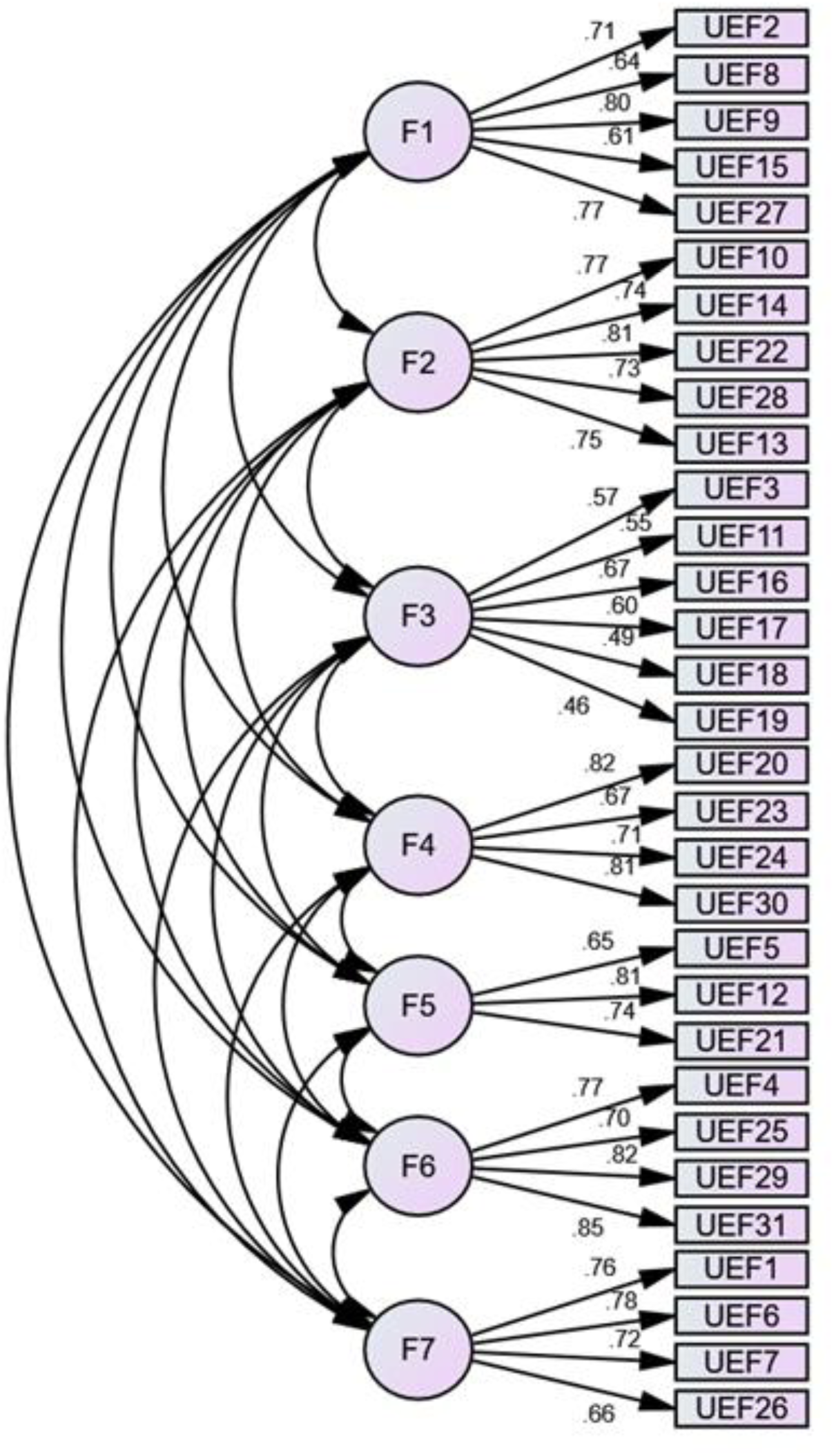
Confirmatory F Analysis of the Executive Function Scale in Cuban University Students (UFE-C).

Additionally, reliability estimates were high for the total score (α = .91, ω = .91). Regarding factor analysis, there were predominantly medium values in conscious monitoring of responsibilities (α = .76, ω = .77), supervisory attention system (α = .83, ω = .83), behavior verification for learning (α = .76, ω = .76), decision making (α = .70, ω = .70), conscious regulation of emotions (α = .82, ω = .82), and task-solving element management (α = .76, ω = .76). There were medium-low values for the conscious regulation of behavior dimension (α = .65, ω = .65).

Additionally, normative scores for the executive function scale for the Cuban university population (UEF-C; Table 4) were calculated. These scores allow for contextualizing and better understanding of individual results by comparing them with the average performance of Cuban university students.

**Table 4.**
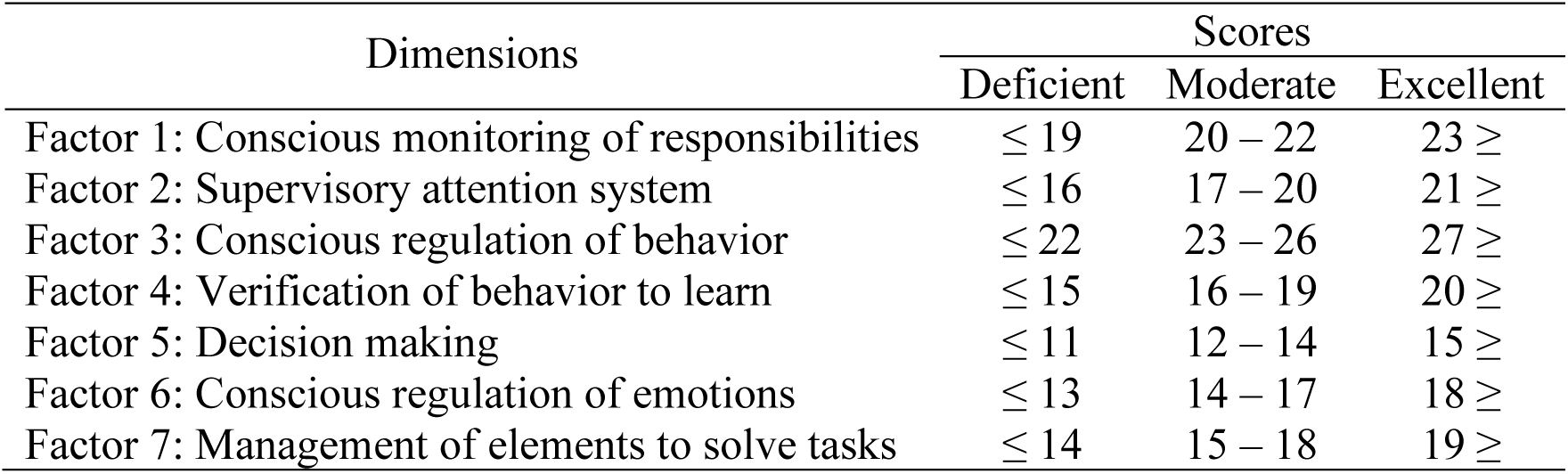
UEF-C scores.

| Dimensions | Scores |  |  |
| --- | --- | --- | --- |
|  | Deficient | Moderate | Excellent |
| Factor 1: Conscious monitoring of responsibilities | $\leq 19$ | 20 – 22 | 23 ≥ |
| Factor 2: Supervisory attention system | $\leq 16$ | 17 – 20 | 21 ≥ |
| Factor 3: Conscious regulation of behavior | $\leq 22$ | 23 – 26 | 27 ≥ |
| Factor 4: Verification of behavior to learn | $\leq 15$ | 16 – 19 | 20 ≥ |
| Factor 5: Decision making | $\leq 11$ | 12 – 14 | 15 ≥ |
| Factor 6: Conscious regulation of emotions | $\leq 13$ | 14 – 17 | 18 ≥ |
| Factor 7: Management of elements to solve tasks | $\leq 14$ | 15 – 18 | 19 ≥ |

## Discussion

The purpose of this research was to evaluate the psychometric properties of the UEF-1 in Cuban university students. The results obtained confirm the factorial structure of the original version composed of seven executive functions (Ramos-Galarza et al., 2023).

This research found an adequate fit of the seven-factor model originally proposed by Ramos-Galarza et al. (2023). The indicators found by these authors (CFI = 0.91, SRMR = 0.04, and RMSEA = 0.04) resemble those obtained in the Cuban validation (RMSEA = .056, CFI = .981; TLI = .979). These fit measures provide evidence of the validity of the model and the scale’s quality in terms of its structure and ability to measure the desired constructs (Finch, 2020; Hox, 2021; Koran, 2020; McNeish & Wolf, 2023).

Similarly to the initial design of the scale, in this research, statistically significant and directly proportional correlations of large magnitudes were found between the evaluated executive functions (r = .30 to .61 in the Cuban population and r = .41 to .70 in the Ecuadorian and Chilean population) (Ramos-Galarza et al., 2023). These scores imply that responses to the different items composing each factor are similar to each other. This suggests that the items are consistently measuring the same construct or dimension (Schober et al., 2018; Tavakol & Wetzel, 2020).

The total reliability estimates of the original scale were similar to those obtained in the Cuban university population. However, the factor estimates in Cuban students showed slightly lower values than those obtained in the original scale (Ramos-Galarza et al., 2023). The main difference was observed in the dimension of conscious regulation of behavior (α = .65, ω = .65) compared to the original sample (α = 0.76 and ω = 0.74). However, all values are within the acceptable range of reliability (Edwards et al., 2021; Fu et al., 2022).

The results obtained in this research are consistent with those obtained by other authors who evaluated executive functions in students (Escolano-Pérez et al., 2022; Kamradt et al., 2019; Prosen & Vitulic, 2014; Ramos-Galarza et al., 2023). All these authors agree that to improve students’ academic performance, daily functioning, and future job prospects, university programs must promote the development of executive functioning skills. This involves providing students with knowledge about these skills and supporting their efficient use.

The present study has relevant implications regarding the assessment of executive functions in real-life situations of Cuban university students. First, the availability of the UEF-C as a reliable and valid instrument for the Cuban context provides a valuable tool for assessing executive functions in real-life situations of Cuban university students. This instrument can facilitate the profiling of performance, inform the development of tailored neuropsychological interventions, and contribute to enhancing students’ executive function skills, ultimately improving their academic and daily functioning. Additionally, our study replicates the findings from the original study, a critical aspect of ensuring that (neuro)psychological measures are applicable and meaningful across different cultural contexts. In this sense, our results play a crucial role in advancing our knowledge of cross-cultural variations in psychological constructs and ensuring the validity of psychological measures across diverse populations.

Despite the contributions of this study, several limitations warrant consideration. The exclusive use of self-reported data, the reliance on predominantly student samples, and the use of snowball sampling for participant recruitment may limit the generalizability of our findings. Future research should aim to address these limitations by incorporating diverse samples, employing objective measures of executive functions, and exploring the nomological validity and temporal stability of the UEF-1 scores over time.

## Conclusions

In conclusion, this study provides valuable insights into the psychometric properties of the UEF-1 in Cuban university students, confirming its validity and reliability as a measure of executive functions in this population. By advancing our understanding of executive functions and their assessment in diverse cultural contexts, this research contributes to the broader field of neuropsychological assessment and intervention, highlighting the importance of promoting executive function skills to support student’s academic success and daily functioning.

## Data Availability

The datasets generated for this study are available on request to the corresponding author

